# Validation of the IMPEDE VTE Score for Prediction of Venous Thromboembolism in Multiple Myeloma: A Retrospective Cohort Study

**DOI:** 10.1101/2020.12.10.20247320

**Authors:** Fahrettin Covut, Ramsha Ahmed, Sanchit Chawla, Frank Ricaurte, Christy J. Samaras, Faiz Anwer, Alex V. Mejia Garcia, Dana E Angelini, Sandra Mazzoni, Beth Faiman, Jason Valent, Jack Khouri

## Abstract

Venous thromboembolism (VTE) is highly prevalent in Multiple Myeloma (MM) patients, however a reliable VTE prediction tool in MM remains under study. The IMPEDE VTE score has recently emerged as a novel risk prediction tool for VTE in MM but needs external validation in different cohorts. We conducted a retrospective cohort study to validate this score. We reviewed 839 patients who were newly diagnosed with MM between 2010 and 2015 at Cleveland Clinic and included 575 patients in final analysis. The 6-month cumulative incidence of VTE among all patients was 10.7% (95% CI: 8.2 – 13.2) and the c-statistic of the IMPEDE VTE score to predict VTE within 6 months of treatment start was 0.68 (95% CI: 0.61 – 0.75). The 6-month cumulative incidence of VTE was 5.0% (95% CI: 2.1 – 7.9) in the low risk group, compared to 12.6% (95% CI: 8.9% – 16.4%) and 24.1% (95% CI: 12.2 – 36.1) in the intermediate and high risk groups (p<0.001 for both). In addition, a higher proportion of patients in the VTE cohort had ECOG performance status of ≥2 as compared to the no VTE cohort (33% vs. 16%, p=0.001). Other MM characteristics such as stage, immunoglobulin subtype, and cytogenetics were not found to be predictors of VTE. In summary, we have validated the IMPEDE VTE score in our patient cohort and our findings suggest that it can be utilized as a VTE risk stratification tool in prospective studies looking into investigating VTE prophylaxis strategies in MM patients.

## BACKGROUND

Venous thromboembolism (VTE) is an important cause of morbidity and mortality in cancer patients ^1,2^. Multiple Myeloma (MM) is the second most common hematological malignancy and associated with 9-fold increased risk of VTE compared to the general population ^3,4^. The higher risk of VTE in MM patients is multifactorial and related to patient-, MM- and treatment-related factors ^5^. The monoclonal protein can lead to hyperviscosity in 2% to 6% of MM patients ^6^, interfere with fibrin clot formation ^7^, and act as a procoagulant antibody ^8^. Elevated cytokine levels in MM patients such as interleukin-6, tumor necrosis factor alpha, and vascular endothelial growth factor further enhance the thrombosis risk ^9^. Immobility due to increased incidence of bone fractures ^10^, use of central venous catheter ^11^, and erythropoiesis-stimulating agents ^12^ are additional risk factors for VTE in MM. Several commonly used plasma cell directed drugs, such as dexamethasone, immunomodulatory drugs (IMiD), alkylating agents, and doxorubicin, alter hemostatic pathways and thus promote thrombogenesis ^13^.

A majority of VTE events in MM patients occur within 6 months of treatment initiation ^14^. In 2008, the International Myeloma Working Group (IMWG) recommended different VTE prophylaxis strategies based on a number of VTE risk factors; aspirin and low molecular weight heparin/warfarin were offered for patients with ≤1 and ≥2 risk factors, respectively ^15^. These recommendations were based on consensus opinion and limited evidence ^15^. Despite the implementation of those guidelines in the Myeloma XI trial, a phase III multicenter randomized clinical trial which included 4358 newly diagnosed MM patients, the cumulative incidence of VTE within 6 months of induction therapy was more than 10% in all study arms ^14^. Importantly, only 55% of patients who had thrombotic events in this trial were in the high-risk group for VTE based on the IMWG criteria ^14^. Sanfilippo et al. have recently developed the IMPEDE VTE score, a risk prediction tool for VTE in MM, using 4446 patients within the veterans administration central cancer registry and validated this using 4256 patients in the Surveillance, Epidemiology, End Results (SEER)-Medicare database ^16^. Patients were stratified into three different risk groups based on the IMPEDE VTE score and the 6-month cumulative incidence of VTE were 15.2%, 8.3%, and 3.3% among patients in the high-risk, intermediate-risk, and low-risk groups at the time of induction, respectively ^16^.

Therefore, the IMPEDE VTE score may provide a better risk stratification for VTE in newly diagnosed MM patients compared to current risk stratification strategies. Herein, we validated the IMPEDE VTE score in our patient cohort.

## METHODS

### Study design and patients

In this cohort study, we retrospectively reviewed all patients who were newly diagnosed with MM at Cleveland Clinic between January 2010 and August 2015. We included patients older than 18 years and received their induction therapy at our institution. We excluded patients who had another active malignancy at diagnosis, did not receive plasma cell-directed therapy, had missing baseline data for calculation of the IMPEDE VTE score, were lost to follow-up within 6 months of treatment initiation and were diagnosed with VTE after MM diagnosis but before initiation of systemic therapy. This study was conducted according to the Declaration of Helsinki and approved by the institutional review board of Cleveland Clinic.

### Study variables and assessments

We retrospectively collected the following data from electronic medical records of the patients; age, gender, race, body mass index (BMI), details of VTE, recent surgeries and fractures, performance status, MM subtype, prognostic stage, cytogenetics, extramedullary disease, laboratory values at diagnosis, use of erythropoietin or central venous catheter, induction regimens, treatment responses, antiplatelet and anticoagulant agents, autologous hematopoietic cell transplantation (AHCT) status, and survival data.

The IMPEDE VTE score at treatment initiation was calculated according to the following parameters: IMID use (+4 points), BMI ≥25 kg/m^2^ (+1 point), recent pelvic, hip, or femur fracture (+4 points), erythropoiesis-stimulating agent (+1 point), doxorubicin (+3 points), dexamethasone (+4 and +2 points for high- and low-dose, respectively), Asian/pacific islander race (−3 points), prior history of VTE (+5 points), central venous catheter (+2 points), therapeutic low molecular weight heparin or warfarin use (−4 points), prophylactic low molecular weight or aspirin use (−3 points) ^16^. Patients with IMPEDE VTE scores of ≤3, 4-7, and ≥8 were considered to have low risk, intermediate risk, and high risk for developing VTE within 6 months of treatment initiation^16^.

High risk cytogenetics were defined as presence of del(13) on karyotype, t(4;14), t(14;16), t(14;20), del(17/17p), or gain(1q) ^17^. Recent surgery was defined as cardiovascular, orthopedic, abdominal, urologic, or neurologic surgery that occurred between 30 days prior to MM diagnosis and the start of systemic therapy ^16^. Similarly, a recent fracture was defined as pelvic, femur or hip fractures occurring between 30?days prior to MM diagnosis and the start of systemic therapy ^16^. If multiple baseline laboratory and BMI data were available, we recorded the values closest to the date of treatment initiation. High-dose and low-dose dexamethasone were defined as the total monthly dose of ≥160 mg and <160 mg, respectively ^18^. Prophylactic and therapeutic enoxaparin doses were defined as 40 mg daily and 1 mg/kg twice daily, respectively ^19,20^. In our practice, patients received VTE prophylaxis according to the IMWG guidelines ^15^, unless bleeding risk outweighed the benefit of VTE prophylaxis or patient preference otherwise. Response to first line of induction treatment was determined based on the IMWG criteria ^21^.

### Statistical Analysis

No sample size calculations were performed. Descriptive analysis was performed to compare baseline characteristics of patients who were and were not diagnosed VTE within 6 months of treatment initiation. Continuous variables were reported as median with interquartile range (IQR) and medians of two groups were compared using Wilcoxon-Mann-Whitney test. Categorical variables were compared using chi–square or Fisher’s exact test. Receiver Operating Characteristic curve analysis was performed and reported as c-statistic to assess the discriminative power of IMPEDE VTE score to predict VTE within 6 months of treatment start. Cumulative incidence of VTE was calculated using Kaplan–Meier method, with death as competing risk, and compared using Gray’s test ^22^. Patients who underwent AHCT were censored at the time of transplant while calculating cumulative incidence of VTE. Overall survival (OS) and progression-free survival (PFS) were calculated from date of MM diagnosis. Multivariable Cox hazard analysis was performed to evaluate the effect of VTE within 6 months of treatment start on PFS and OS. All reported p values are two–sided and a p value of less than 0.05 was considered statistically significant. All statistical calculations were made using R statistical software version 3·4·0 (R Foundation for Statistical Computing, Vienna, Austria).

## RESULTS

### Baseline Characteristics of the Patients

We reviewed the electronic medical records of 839 MM patients, excluded 264 of them, and analyzed the final cohort of 575 patients. Figure 1 summarizes the patient selection process. Two hundred eighty-eight (50%) patients were male, 456 (79%) were white, 258 (45%) aged >65 years, and 409 (71%) had BMI of ≥ 25 kg/m^2^. Median age and BMI at diagnosis were 64 years (IQR: 56 – 71) and 28.1 kg/m^2^ (IQR: 24.4 – 32.6). Table 1 summarizes the baseline characteristics of 61 (10.6%) patients who developed VTE (VTE cohort) and 514 (89.4%) patients who did not develop VTE within 6 months upon treatment initiation (no VTE cohort).

**Figure 1.**
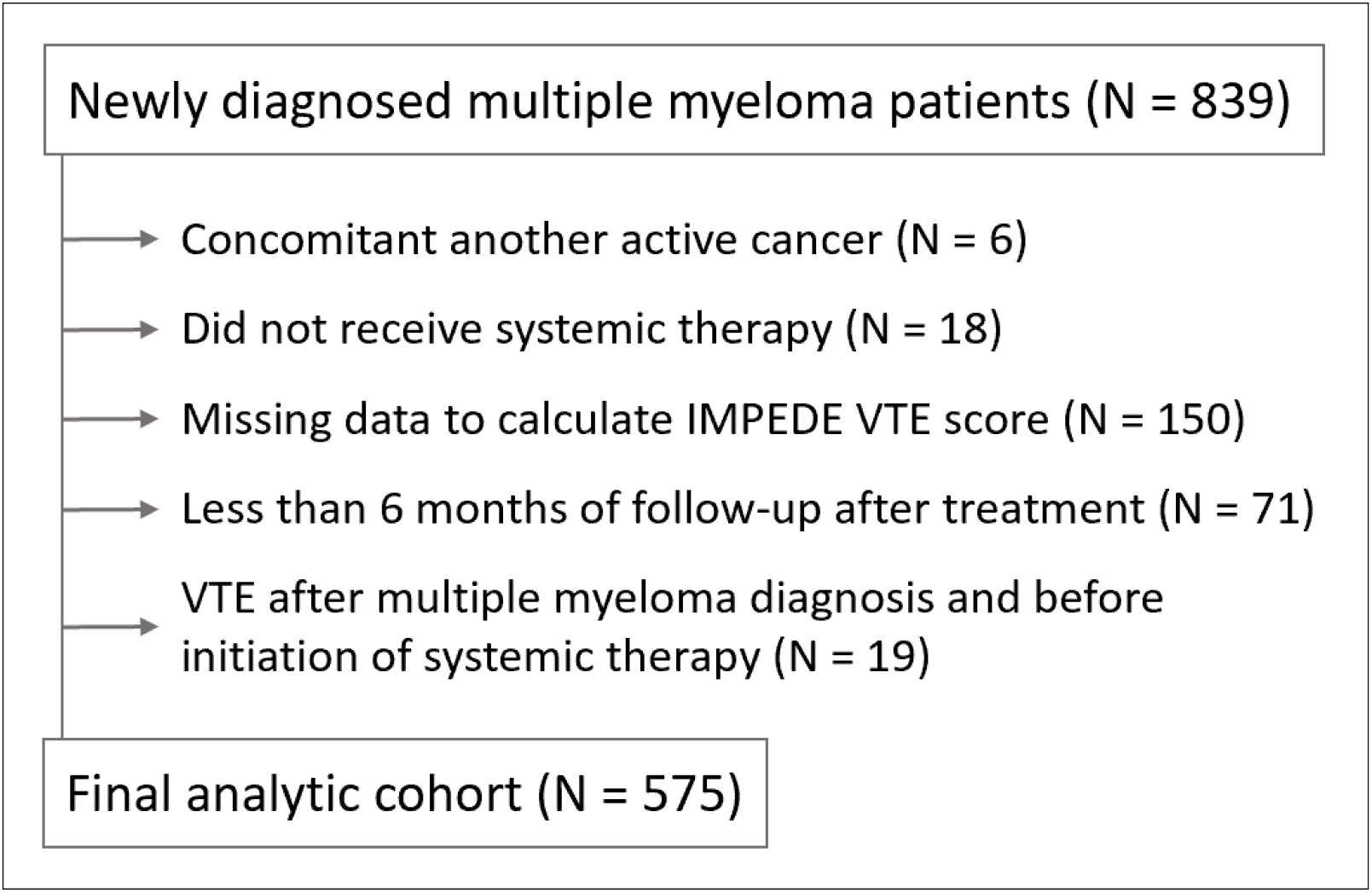
Flow diagram of patient selection process.

**Table 1.** Baseline characteristics of the patients.

| Characteristics | All patients<br>(N = 575) | VTE<br>in 6 months<br>(N = 61) | No VTE<br>in 6 months<br>(N = 514) | P value |
| --- | --- | --- | --- | --- |
| Age, year – median (IQR) | 63.6 (56.4 – 71.2) | 65.4 (57.3 – 71.5) | 63.5 (56.4 – 71.2) | 0.14 |
| Age > 65 years – no. (%) | 258 (44.9) | 31 (50.8) | 227 (44.2) | 0.32 |
| Male gender – no. (%) | 288 (50.1) | 32 (52.5) | 256 (49.8) | 0.70 |
| Race – no. (%) |  |  |  | 0.82 |
| White | 456 (79.3) | 48 (78.7) | 408 (79.4) |  |
| Black | 116 (20.2) | 13 (21.3) | 103 (20.0) |  |
| Asian / pacific islander | 3 (0.5) | 0 (0) | 3 (0.6) |  |
| Body mass index, kg/m<sup>2</sup> – median (IQR) | 28.1 (24.4 – 32.6) | 30.4 (26.7 – 34.0) | 28.0 (24.4 – 32.3) | 0.052 |
| Body mass index ≥ 25 kg/m<sup>2</sup> – no. (%) | 409 (71.1) | 47 (77.0) | 362 (70.4) | 0.29 |
| History of venous thromboembolism – no. (%) | 31 (5.4) | 10 (16.4) | 21 (4.1) | <0.001 |
| Pathologic pelvic or femur fracture – no. (%) | 16 (2.8) | 6 (9.8) | 10 (1.9) | <0.001 |
| Surgery – no. (%) | 19 (3.3) | 6 (9.8) | 13 (2.5) | 0.003 |
| ECOG performance status – no. (%) |  |  |  | 0.001 |
| < 2 | 475 (82.6) | 41 (67.2) | 434 (84.4) |  |
| ≥ 2 | 100 (17.4) | 20 (32.8) | 80 (15.6) |  |
| Subtype of multiple myeloma – no. (%) |  |  |  | 0.77 |
| IgG | 311 (54.1) | 34 (55.7) | 277 (53.9) |  |
| IgA | 122 (21.2) | 10 (16.4) | 112 (21.8) |  |
| Light chain disease | 128 (22.3) | 15 (24.6) | 113 (22.0) |  |
| IgD, IgM, or Asecreatory disease | 14 (2.4) | 2 (3.3) | 12 (2.3) |  |
| International staging system – no. (%) |  |  |  | 0.08 |
| I | 182 (31.7) | 26 (42.6) | 156 (30.4) |  |
| II | 177 (30.8) | 16 (26.2) | 161 (31.3) |  |
| III | 175 (30.4) | 13 (21.3) | 162 (31.5) |  |
| Extramedullary disease – no. (%) | 151 (26.3) | 18 (29.5) | 133 (25.9) | 0.54 |
| Cytogenetics at diagnosis * – no. (%) |  |  |  |  |
| Del(13) on karyotype | 42 (7.3) | 7 (11.5) | 35 (6.8) | 0.19 |
| Del(17/17p) | 35 (6.1) | 4 (6.6) | 31 (6.0) | 0.87 |
| t(4;14) | 46 (8.0) | 3 (4.9) | 43 (8.4) | 0.73 |
| t(11;14) | 109 (19.0) | 16 (26.2) | 93 (18.1) | 0.13 |
| t(14;16) | 22 (3.8) | 2 (3.3) | 20 (3.9) | 0.81 |
| t(14;20) | 2 (0.3) | 1 (1.6) | 1 (0.2) | 0.20 |
| Gain(1q) | 25 (4.3) | 3 (4.9) | 22 (4.3) | 0.82 |
| Laboratory tests at diagnosis – median (IQR) |  |  |  |  |
| White blood cell count, 10<sup>6</sup>/uL | 5.8 (4.5 – 7.5) | 5.9 (4.5 – 7.2) | 5.8 (4.5 – 7.5) | 0.61 |
| Hemoglobin, g/dL | 10.8 (9.1 – 12.1) | 10.9 (9.3 – 11.9) | 10.8 (9.0 – 12.2) | 0.93 |
| Platelet count, 10<sup>9</sup>/uL | 211 (158 – 268) | 219 (168 – 271) | 210 (155 – 267) | 0.66 |
| Beta-2-microglobulin, mg/L | 3.8 (2.6 – 6.4) | 3.2 (2.6 – 5.4) | 3.9 (2.6 – 6.4) | 0.11 |
| Albumin, gm/dL | 3.7 (3.2 – 4.1) | 3.7 (3.2 – 4.2) | 3.7 (3.2 – 4.1) | 0.78 |
| Creatinine, mg/dL | 1.0 (0.8 – 1.5) | 1.0 (0.8 – 1.4) | 1.0 (0.8 – 1.5) | 0.75 |
| Lactate dehydrogenase, U/L | 180 (150 – 220) | 192 (172 – 239) | 178 (149 – 219) | 0.09 |

\* Cytogenetic data were not available for 50 (9%) patients. In addition to karyotype analysis, FISH probes for del(17/17p), t(4;14), t(11;14), t(14;16), t(14;20), and gain(1q) were used in 326 (57%), 186 (32%), 255 (44%), 172 (30%), 24 (4%), and 70 (12%) patients, respectively.

Fifty-one (8.9%), 304 (52.9%), and 220 (38.3%) patients had IMPEDE VTE score of ≥8 (high risk), 4 to 7 (intermediate risk), and ≤3 (low risk) at treatment start. Fourteen (27%) patients were black in high risk group, compared to 67 (22%, p=0.39) and 35 (16%, p=0.05) patients in intermediate and low risk groups per IMPEDE VTE score, respectively. The median IMPEDE VTE score was 4 (IQR: 3 – 6) in all patients and 5 (IQR: 4 – 7) vs. 4 (IQR: 3 – 5) in the VTE vs. no VTE cohorts, respectively (p<0.001). MM patients in the VTE cohort were more likely to have a history of previous VTE (16% vs. 4%, p<0.001), a recent pathologic pelvic, hip, or femur fracture (10% vs. 2%, p<0.001), recent surgery (10% vs. 3%, p=0.003), and ECOG performance status of ≥2 (33% vs. 16%, p=0.001) compared to those in the no VTE cohort.

### Treatment Characteristics and VTE prophylaxis

Table 2 summarizes treatment data. Within 6 months of treatment initiation, there was no difference in the use of central venous catheters (25% vs. 17%, p=0.13), erythropoiesis-stimulating agents (3% vs. 6%, p=0.56), IMID (75% vs. 67%, p=0.17), bortezomib (67% vs. 71%, p=0.58), melphalan (2% vs. 4%, p=0.71), cyclophosphamide (7% vs. 11%, p=0.38), and doxorubicin (0% vs. 1%, p=1) between the VTE and no VTE cohorts, respectively. High-dose dexamethasone was more frequently used in the VTE cohort as compared to the no VTE cohort (15% vs. 2%, p<0.001). Seventy-five percent of patients in the VTE cohort and 72% of patients in the no VTE cohort were on an anti-platelet or anticoagulant agent at treatment start (p=0.55). Five (8%) and 32 (6%) patients in VTE and no VTE cohorts underwent AHCT within 6 months of treatment start (p=0.58).

**Table 2.** Treatment characteristics and outcomes of the patients.

| Characteristics | All patients<br>(N = 575) | VTE<br>in 6 months<br>(N = 61) | No VTE<br>in 6 months<br>(N = 514) | P-value |
| --- | --- | --- | --- | --- |
| Central venous catheter – no. (%) | 101 (17.6) | 15 (24.6) | 86 (16.7) | 0.13 |
| Erythropoiesis-stimulating agent – no. (%) | 34 (5.9) | 2 (3.3) | 32 (6.2) | 0.56 |
| Drugs used within 6 months of treatment start – no. (%) |  |  |  |  |
| Thalidomide | 22 (3.8) | 2 (3.3) | 20 (3.9) | 1 |
| Lenalidomide | 373 (64.9) | 44 (72.1) | 329 (64.0) | 0.21 |
| Pomalidomide | 3 (0.5) | 0 (0) | 3 (0.6) | 1 |
| Bortezomib | 404 (70.3) | 41 (67.2) | 363 (70.6) | 0.58 |
| Melphalan | 19 (3.3) | 1 (1.6) | 18 (3.5) | 0.71 |
| Cyclophosphamide | 61 (10.6) | 4 (6.6) | 57 (11.1) | 0.38 |
| Doxorubicin | 3 (0.5) | 0 (0) | 3 (0.6) | 1 |
| Dexamethasone in initial induction regimen – no. (%) |  |  |  | <0.001 |
| Low-dose dexamethasone | 544 (94.6) | 50 (82.0) | 494 (96.1) |  |
| High-dose dexamethasone | 21 (3.7) | 9 (14.8) | 12 (2.3) |  |
| Not used | 10 (1.7) | 2 (3.3) | 8 (1.6) |  |
| Number of drugs in initial induction regimen – no. (%) |  |  |  | 0.26 |
| Single agent | 6 (1.0) | 2 (3.3) | 4 (0.8) |  |
| Doublet | 335 (58.3) | 34 (55.7) | 301 (58.6) |  |
| Triplet | 223 (38.8) | 23 (37.7) | 200 (38.9) |  |
| Quadruplet | 11 (1.9) | 2 (3.3) | 9 (1.8) |  |
| Anti-platelet or anticoagulant agents – no. (%) |  |  |  |  |
| Not used | 160 (27.8) | 15 (24.6) | 145 (28.2) | 0.55 |
| Aspirin or clopidogrel | 357 (62.1) | 37 (60.7) | 320 (62.3) | 0.81 |
| Warfarin | 40 (7.0) | 6 (9.8) | 34 (6.6) | 0.35 |
| Prophylactic enoxaparin | 15 (2.6) | 3 (4.9) | 12 (2.3) | 0.21 |
| Direct oral anticoagulants | 3 (0.5) | 0 (0) | 3 (0.6) | 1 |
| Best response to initial induction regimen – no. (%) |  |  |  | 0.89 |
| Very good partial response or better | 266 (46.3) | 28 (45.9) | 238 (46.3) |  |
| Partial response | 171 (29.7) | 20 (32.8) | 151 (29.4) |  |
| Stable or progressive disease | 110 (19.1) | 11 (18.0) | 99 (19.3) |  |
| Unknown | 28 (4.9) | 2 (3.3) | 26 (5.1) |  |
| Autologous hematopoietic cell transplantation – no. (%) | 202 (35.1) | 17 (27.9) | 185 (36.0) | 0.21 |
| < 6 months after treatment initiation | 37 (6.4) | 5 (8.2) | 32 (6.2) | 0.58 |

### Outcomes

The 6-month cumulative incidence of VTE among all patients was 10.7% (95% CI: 8.2 – 13.2). Each one point increase in the IMPEDE VTE score was associated with VTE within 6 months of treatment start (Cox hazard ratio: 1.25, 95% CI: 1.14 – 1.36, p<0.001). The c-statistic of the IMPEDE VTE score to predict VTE within 6 months of treatment start was 0.68 (95% CI: 0.61 – 0.75). The 6-month cumulative incidence of VTE according to the IMPEDE VTE score was 5.0% (95% CI: 2.1 – 7.9) in the low risk group, compared to 12.6% (95% CI: 8.9% – 16.4%) and 24.1% (95% CI: 12.2 – 36.1) in the intermediate and high risk groups (p<0.001 for both), respectively (Figure 2). The median follow-up of patients was 60.3 months (IQR: 25.9 – 78.5). On multivariable Cox hazard analysis, VTE within 6 months of treatment initiation was not a predictor of PFS (HR: 1.13, 95% CI: 0.80 – 1.61, p=0.49) and OS (HR: 1.31, 95% CI: 0.85 – 2.04, p=0.23) after adjusted for age, performance status, prognostic stage, cytogenetics, number of drugs used in initial induction, and AHCT (Table 3).

**Table 3.** Multivariable Cox hazard analysis for progression-free and overall survival.

| Variables | Progression-free survival |  | Overall survival |  |
| --- | --- | --- | --- | --- |
|  | Hazard ratio (95% CI) | P value | Hazard ratio (95% CI) | P value |
| Age at diagnosis |  |  |  |  |
| Each 1-year increase | 1.01 (1.00 – 1.02) | 0.12 | 1.02 (1.00 – 1.03) | 0.025 |
| ECOG performance status |  |  |  |  |
| Each 1 increase | 1.32 (1.17 – 1.50) | <0.0001 | 1.48 (1.27 – 1.71) | <0.0001 |
| International staging system |  |  |  |  |
| Each increase in risk category | 1.23 (1.07 – 1.42) | 0.003 | 1.52 (1.27 – 1.81) | <0.0001 |
| Cytogenetics |  |  |  |  |
| High risk vs. standard risk | 2.18 (1.70 – 2.79) | <0.0001 | 2.75 (2.06 – 3.66) | <0.0001 |
| Number of drugs in initial induction regimen |  |  |  |  |
| Each 1 increase | 0.79 (0.65 – 0.96) | 0.020 | 0.65 (0.50 – 0.84) | 0.0008 |
| Autologous hematopoietic cell transplantation |  |  |  |  |
| Performed vs. not performed | 0.73 (0.57 – 0.94) | 0.014 | 0.62 (0.44 – 0.88) | 0.007 |
| VTE within 6 months of treatment start |  |  |  |  |
| Yes vs. no | 1.13 (0.80 – 1.61) | 0.49 | 1.31 (0.85 – 2.04) | 0.23 |

**Figure 2.**
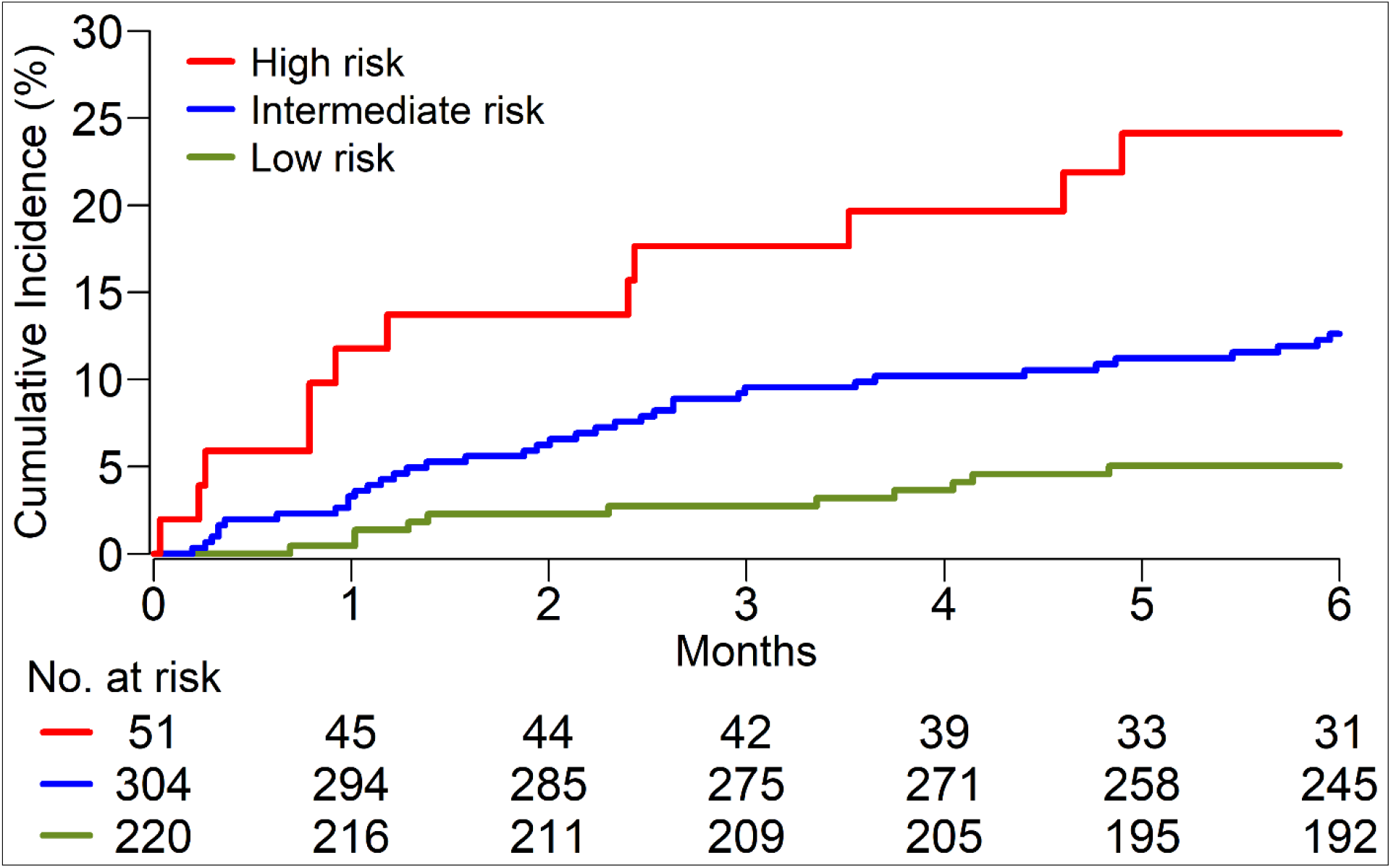
Cumulative incidence of venous thromboembolism after initiation of treatment.

## DISCUSSION

In this large cohort of newly diagnosed MM patients, we show that the IMPEDE VTE score successfully stratified patients into three distinct VTE risk groups. The discriminative power of this score in our cohort was similar to the original study ^16^. Although poor performance status was not incorporated into the IMPEDE VTE score, it was significantly associated with VTE in our cohort. Though 72% of our patients received VTE prophylaxis with either antiplatelet or prophylactic enoxaparin, roughly 1 of 10 patients in the entire cohort were diagnosed with VTE within 6 months of treatment start, similar to the Myeloma XI trial ^14^. The 6-month cumulative incidence of VTE was more than 10% among our patients in the intermediate and high risk groups according to the IMPEDE VTE score. Thus, one may infer that routine VTE prophylaxis is not enough for these patients. We have not identified any association between VTE and MM prognostic stage, immunoglobulin subtype, or cytogenetics. Disparities in outcomes of white and black MM patients were previously noted ^23^. Although we observed higher proportion of blacks in higher risk groups per IMPEDE VTE score, blacks and whites had similar rates of VTE. Patients who developed VTE had similar OS and PFS compared to patients in the no VTE cohort.

The rates of VTE in MM patients receiving therapy with IMiD or high-dose dexamethasone have been reported to be between 20% and 30% without VTE prophylaxis ^18,24^. Although routine VTE prophylaxis is the standard of care for these patients, there are a few prospective trials that have investigated the optimal choice and dose of thromboprophylaxis in this setting. In a phase 3 trial, 667 patients were randomized to receive low dose aspirin, low dose warfarin, or prophylactic enoxaparin for the first 6 months of thalidomide-based therapy and similar rates of thrombotic events, 5% to 8%, were seen in all arms ^25^. This study excluded patients with prior history of VTE and other cardiovascular risk factors ^25^. In another randomized trial, 2.3% and 1.2% of patients who received lenalidomide-based therapy had thrombotic events within first 12 months in low dose aspirin and prophylactic enoxaparin arms, respectively ^26^. Patients with a higher risk of VTE were also excluded from this study, including elderly patients (age >65 years), patients with previous VTE and other high-risk conditions ^26^. The exclusion of patients with strong VTE risk factors from the clinical trials limits the applicability of these results in the real-world setting.

Although the current IMWG guidelines recommend warfarin or low-molecular-weight-heparin for patients at a higher risk of VTE, direct oral anticoagulants (DOACs) have also been found to be safe and effective in cancer patients. In 2019, two randomized controlled trials have assessed the utility of DOACs for prevention of VTE among ambulatory cancer patients undergoing chemotherapy. In the AVERT trial, apixaban was associated with a significantly lower incidence of VTE compared to placebo ^27^. In the CASSINI trial, rivaroxaban was associated with a significantly lower incidence of VTE per-protocol analysis but not per intention-to-treat analysis compared to placebo ^28^. Cumulative analysis of both trials has shown significant benefit of DOACs for the prevention of VTE, with a low incidence of major bleeding ^29^. These trials used the Khorana score for VTE risk stratification which does not apply to MM patients ^30^. Two trials have evaluated DOACs for VTE prophylaxis in MM patients. In a multicenter, non-randomized, phase 2 trial, 104 patients with newly diagnosed or relapsed MM received apixaban 2.5 mg twice daily during the first 6 months of IMiD-based therapy; only 2 patients had DVT and 1 patient had major bleeding ^31^. In a prospective proof-of-concept study, 50 MM patients received apixaban 2.5 mg twice daily for 6 months while receiving IMiD-based induction or maintenance therapy, and none of them had DVT or major bleeding ^32^. Overall, the safety and efficacy data in single arm trials using DOACs for VTE prophylaxis in MM patients look promising, but further prospective data is needed. A prospective randomized controlled trial of apixaban 2.5 mg twice daily vs. placebo in MM patients receiving IMiD-based therapy is ongoing (NCT02958969). The IMPEDE VTE score may be a useful risk stratification tool for future trials investigating the role of DOACs in MM patients.

To our knowledge, this is the first study that independently validates the IMPEDE VTE score in a large cohort of MM patients. We have shown that a poor performance status was associated with a higher incidence of VTE which, if considered, may strengthen the predictive power of the IMPEDE VTE score. Furthermore, despite following current IMWG VTE prophylaxis recommendations, patients in our cohort still had an VTE rate of >10%, suggesting these current strategies are not enough in this patient population. Some of the limitations of this study include its retrospective design and the exclusion of patients with missing baseline data or shorter follow-up which may have caused selection bias. Extrapolation of our findings may also be limited by the fact that the studied patient cohort was from a single center.

In summary, our findings suggest that the IMPEVE VTE score is a valid evidence-based risk stratification tool in newly diagnosed MM patients with potential to be implemented in future studies aimed at refining our approach to VTE prophylaxis in this patient population. Further validation is required in prospective studies.

## Data Availability

Access to data can be granted on request to qualified investigators for appropriate use, in accordance with the institutional policy to protect the confidentiality of patients.

